# Ensemble Machine Learning for Malaria Diagnosis in Resource-Limited Settings Using Clinical and Demographic Features

**DOI:** 10.1101/2025.08.03.25332923

**Authors:** Panashe Nyengera, Hilary Takawira, Farai Mlambo

## Abstract

**Background:** Sub-Saharan Africa continues to shoulder the heaviest burden of malaria. The 2024 WHO malaria report highlighted that Africa contributed an alarming 94% of the global cases and 95% of the deaths. In the WHO African region, progress towards elimination and management of malaria is hindered by weak health systems, and lack of traditional diagnostic methods such as microscopy and malaria rapid diagnostic tests (mRDT). The primary aim of the study is to develop a machine learning (ML) ensemble model for malaria diagnosis using clinical and demographic data, tailored for resource-limited settings.

**Methods:** A retrospective study was conducted using 637 patient records from Gutu Mission Hospital and Gweru Provincial Hospital in Zimbabwe. Clinical symptoms (fever, chills, abdominal pain, headache and diarrhea) and demographic features (age, gender, residence and travel history) were analysed. Data preprocessing included handling class imbalance using Synthetic Minority Oversampling Technique (SMOTE) and feature selection using Recursive feature elimination (RFE). Seven individual ML models including Logistic regression (LR), Random Forest (RF), Decision Trees (DT), Gradient Boosting (GB), K-Nearest Neighbor (KNN), Naive Bayes (NB) and XGBoost were trained and evaluated on the malaria dataset. The individual models were further combined to build, train and evaluate ensemble models such as Bagging, Stacking, Soft Voting and AdaBoost. Model performance was assessed using accuracy, precision, confusion matrices, recall and F1score and AUR-ROC metrics.

**Results:** Clinical symptoms (chills: p=0.001, fever: p=0.003, diarrhoea: p=0.01, abdominal pain: p<0.001) were statistically significant predictors of malaria. Of the demographic factors, only travel history (p=0.02) showed significant association with malaria. Among the seven individual ML models, GB achieved the highest predictive performance (Accuracy = 0.94), followed by RF (Accuracy = 0.94%) and XGBoost (Accuracy = 0.93%). The stacking ensemble model outperformed all individual ML models and other ensemble models (bagging, soft voting and adaBoost) achieving accuracy = 0.96, precision = 0.95, recall = 0.98, F1 score= 0.96 and AUC-ROC = 0.98.

**Conclusion:** This study demonstrates that ML particularly ensemble models can be used to significantly improve malaria diagnosis. The integration of these models into a web-based application could provide a scalable and accessible diagnostic tool for healthcare workers in resource limited settings.

## BACKGROUND

More than half of the world population is affected by malaria, a parasitic disease that remains a major public health problem (Hamid *et al*., 2024). The 2024 World Health Organization (WHO) malaria report estimates that 263 million cases with 597 000 deaths occurred in 2023. The malaria report also highlights that the WHO African regions, especially Sub-Saharan Africa, continues to shoulder the heaviest burden, contributing 94% of the cases and 95% of the deaths globally (WHO, 2024). Malaria disproportionately affects the rural areas, especially poor communities with limited to no access to healthcare (Rajab *et al*., 2024). Annually, over five million people are at risk of contracting malaria in Zimbabwe. Zimbabwe malaria statistical reports indicated 16 794 cases and 32 deaths in the first half of 2024 (Moyo-Ndlovu, 2024). Of those cases, 199 were children under five years of age, an indication of the persistent burden of malaria within the country. Many at-risk groups continue to miss out on needed services to prevent, diagnose and treat malaria (WHO, 2024).

Malaria is an infectious disease caused by the plasmodium parasite (Kabalu *et al*., 2024). The most severe malaria cases are caused by the *Plasmodium falciparum* species (Wang *et al*., 2019). Traditional parasitology diagnostic methods such as malaria Rapid Diagnostic Tests (mRDTs) and microscopy are widely used for malaria diagnosis (Mfuh *et al*., 2019). Microscopy is the gold standard for malaria diagnosis since the early 20th century but it is time-intensive, requires skilled personnel, and is inclined to variability in accuracy depending on operator expertise (Kabalu *et al*., 2024, Mfu *et al*., 2019). Similarly, mRDTs are one of the most efficient tools used to accurately determine the malaria status of a patient, but its sensitivity is reduced at low parasite densities producing false-negative or false-positive results (Mfu *et al*., 2019). Resource limited settings are usually affected by the absence of a definitive diagnosis, which is a serious obstacle to treatment compliance, efficacy, and clinical care of severe malaria cases (Wang *et al*., 2019).

Traditional statistical methods such as logistic regression, linear regression, and time series analysis, have been used in malaria research to identify risk factors and model malaria prevalence. However, they have been found to depend on data assumptions, focus on explanation over prediction, and they struggle with high-dimensional clinical data but are very useful in understanding malaria trends (Stephen *et al*., 2021). Machine Learning (ML), a subfield of AL that focuses on developing the algorithms that can learn patterns and relationships from data without being explicitly programmed, offers an alternative to supporting malaria diagnosis in resource limited settings.

A study focusing on ML for malaria prediction using clinical and demographic features was carried out in Uganda (Rajab *et al*., 2024). Individual classifiers such as random forest, support vector machines, gradient boosting, decision trees, naive bayes and K-Nearest Neighbors were used to build models using the clinical and demographic features (Rajab *et al*., 2024). The individual ML model performance was evaluated using metrics such as accuracy, precision and recall and they all achieved a good performance (Rajab *et al*., 2024). Ensemble models such as bagging, adaBoost, soft and hard voting and stacking were created from the individual models. The ensemble models outperformed the individual classifiers, achieving over 0.98 across the same metrics (Rajab *et al*. 2024).

In Yumman Province China, a study compared the performance of traditional time series models and deep learning algorithms for malaria prediction (Wang *et al*., 2019). The autoregressive integrated moving average (ARIMA), seasonal and trend decomposition using Loess (STL+ARIMA), back propagation artificial neural network (BP-ANN), and long short-term memory (LSTM) network models were applied separately in simulations using malaria data and meteorological data (Wang *et al*., 2019). GB regression trees were used to combine the four models (ARIMA, STL+ARIMA, BP-ANN and LSTM) through stacking (Wang *et al*., 2019). The findings reported that predictive performance of the stacking ensemble model was superior to that of the individual models, indicating that stacking may have significant implications for malaria disease prediction (Wang *et al*., 2019).

The application of ML based diagnostic models in Zimbabwe remains underexplored. Up till now, limited research has been done to improve the accuracy of malaria diagnosis using ML models. This study aims to bridge this gap by developing an ML ensemble model for malaria diagnosis using readily available clinical and demographic data relevant to malaria. Specific focus will be on adopting ensemble model techniques as they offer a more comprehensive approach. The use of clinical and demographic data for this ensemble model caters for the issue relating to data availability and applicability in underserved communities.

## MATERIALS AND METHODS

### Study design and data collection

Our study followed the methodology described in Figure 1. A retrospective quantitative study was conducted using anonymized patient records from Gweru Provincial Hospital (urban setting) and Gutu Mission Hospital (rural setting) in Zimbabwe, selected to represent high and low malaria transmission areas.

**Figure 1:**
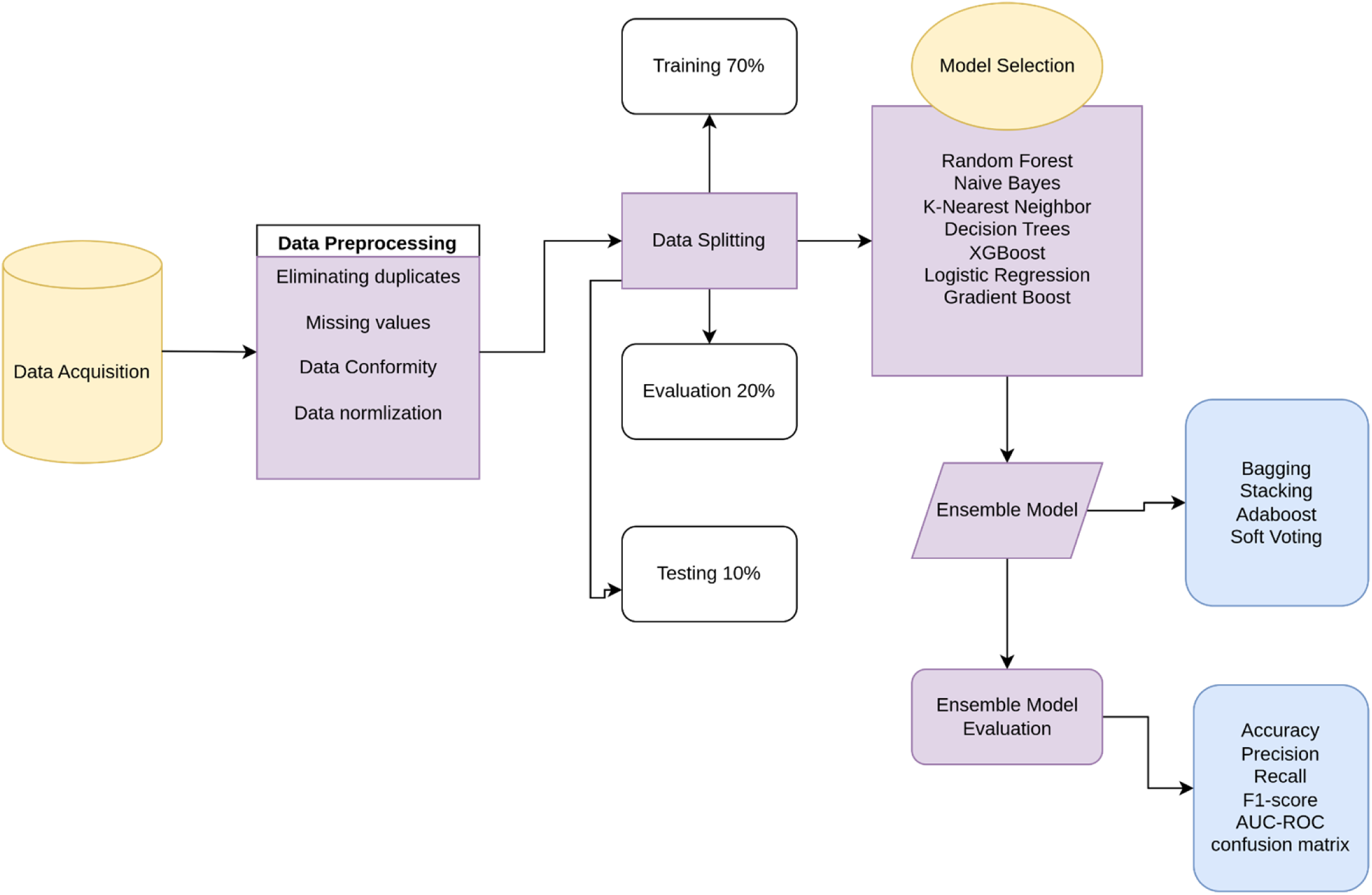
Methodology framework summarizing the various steps of the study from data acquisition up to model evaluation

### Inclusion and exclusion criteria

Patient records from 2022 to December 2024 with a confirmed malaria diagnosis (positive/negative) either through microscopy or mRDTs, with comprehensive clinical and demographic data were included in this study. However, records with missing key variables such as malaria test results, cases with co-infections such as typhoid, or ambiguous diagnostic outcomes were excluded from the study.

### Sample size and sampling procedure

The study used the entire dataset of eligible records, 637 participants tested for malaria with complete details from Gutu Mission Hospital and Gweru Provincial Hospital. The dataset consisted of participants who tested either positive and negative for malaria using microscopy or mRDTs.

### Dependent and independent variables

The outcome variable for this study referred to as malaria status, a binary variable, captures whether the patient was positive or negative for malaria. The predictors included clinical symptoms (fever, chills, headache, abdominal pain, diarrhea (binary variables) and demographic features (age, gender, residence and travel history).

### Data preprocessing

We created a microsoft excel document for the malaria dataset to simplify data pre-processing in R-4.5.1. Data preprocessing in this study included data cleaning, feature selection, handling class imbalance, data encoding, and data split.

### Data Cleaning

Data entry typos, wrong numerical values and incorrect data formats were identified by comparing the paper based medical records and the computed spreadsheet and they were corrected to align with original dataset. Duplicate records were checked for by verifying key identifiers such as patient ID and test date, and were removed to avoid redundant data from skewing the ML models. The dataset was then checked for outliers to make sure no extreme values might affect predictive accuracy. Data points with extreme values were capped, which means replacing extreme values with upper and lower threshold values.

### Feature Selection

Variance inflation factor (VIF) and Pairwise Pearson correlation coefficients were used to study the relationships between the variables. Recursive Feature Elimination (RFE) guided by RF was used for feature selection, choosing the most effective features for model building.

### Handling class imbalance

An imbalanced dataset is when one class outnumbers the other. This imbalance can lead to biases towards the majority class (Basant *et al*., 2024). The majority of the malaria diagnosis results from the collected data were negative (562) compared to the 75 positive cases, very underrepresented in a study comprising of 637 participants. SMOTE was used to address data imbalance using the SMOTE function in R. This was done through creating synthetic samples for the minority class by interpolating between existing minority class.

### Data encoding

One-Hot encoding was used to change categorical variables into numerical representations to make them compatible with ML algorithms. Age was grouped into categorical bins (0-5 years, 6-15 years, 16-30 years, 31-45 years, 46-60 and 61+ years) [Table 1]. The decision to categorize age was based on clinical and epidemiological literature which have successfully proven that working with age bins for malaria related studies takes into consideration that malaria incidence and severity vary non-linearly across age groups and effectively improves model performance (Ranjha *et al*., 2023; Khagayi *et al*., 2019; Carneiro *et al*., 2010)

**Table 1:**
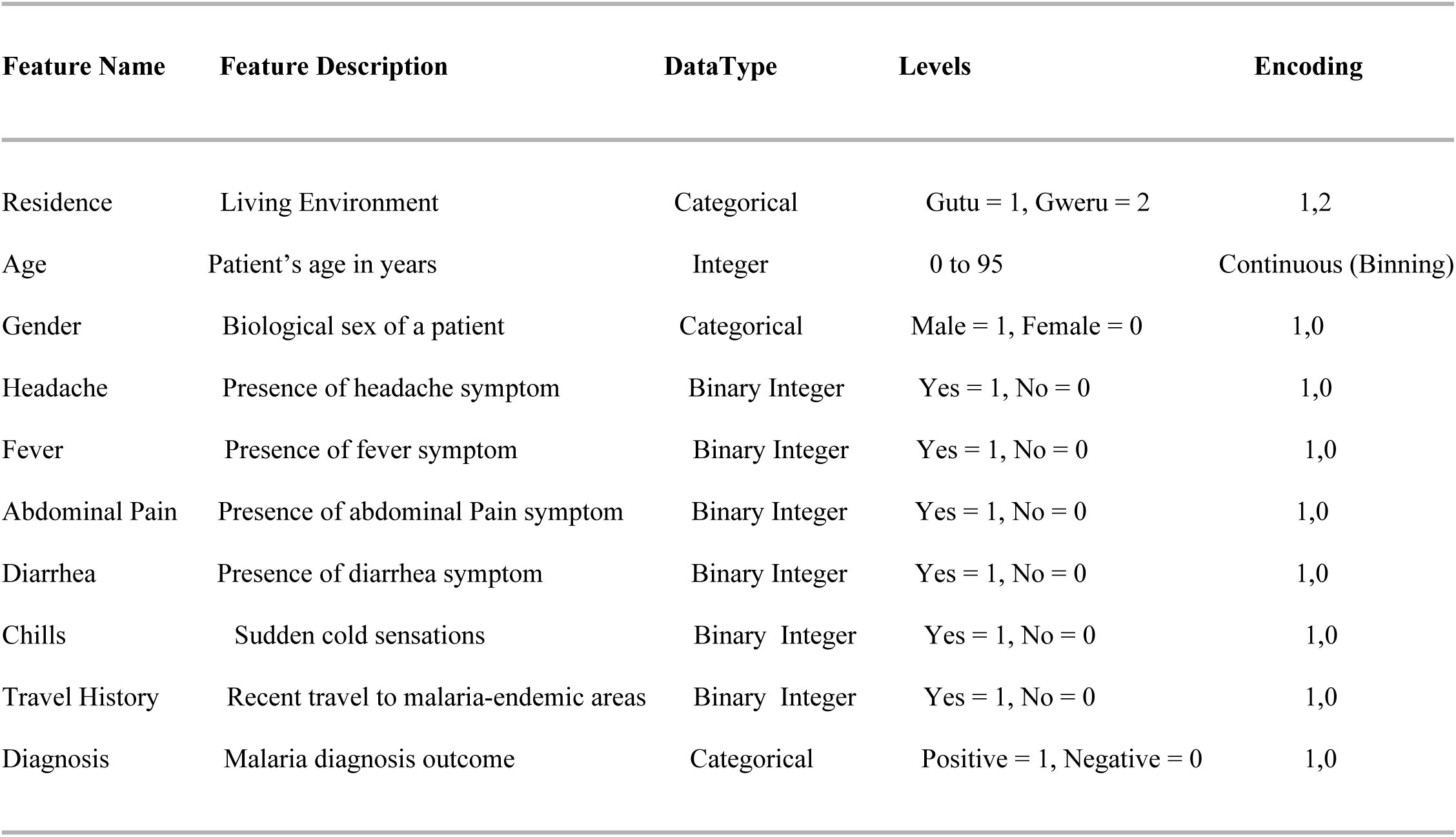
Data Coding Description

### Data splitting

The dataset was split using stratified sampling into training, evaluation and testing sets at a ratio 70:20:10 respectively. The 70% for training allows for sufficient model training and the 20% for evaluation ensures there is enough data for hyperparameter tuning and evaluation of model performance. The 10% for testing was used for final evaluation of model performance and was strictly unseen during training and hyperparameter tuning to provide an unbiased estimate for the model’s performance on unseen data.

### Individual model selection and training

We selected and trained seven individual ML classification algorithms so that we can understand their impact on specific ensemble methods. These models include, LR, GB, KNN, XGBoost, RF, DT and NB. A comprehensive literature review was conducted for selection of predictive classifier models best suited for malaria status prediction (Rajab *et al*., 2024; Muriithi *et al*., 2024; Yadav *et al*., 2021). The seven ML models were trained using 70% of both the balanced and unbalanced datasets to compare how well they performed.

### Hyperparameter tuning

Optimization of model performance was done by tuning hyperparameters relevant to specific ML classifiers. Lasso regression was used to prevent overfitting and improve generalization of LR. LR was also trained on 5 fold cross-validation to improve performance.

RF and XGBoost were tuned using Grid Search. The hyperparameter mtry in RF was tested for values c(1, 2, 3, 4) and 10 fold cross-validation was applied to verify for favourable parameter settings. For XGBoost, the hyperparameters tuned included the number of trees, learning rate and maximum depth. 5 cross-fold validation was used to evaluate the most favorable hyper parameters for XGBoost.

Cross validation based pruning was used for DT. It was applied to optimize the tree’s complexity and prevent overfitting using rpart function in R. Random search was used for tuning hyperparameters for KNN where tune length was set to 10 to allow KNN to explore a range of values for k. For GB, bayesian optimization was used to tune the hyperparameters such as number of trees, learning rate and tree dept. Cross validation was used for NB and tuneLength was set to 10.

### Ensemble model selection and building

Using ensemble techniques through the integration of various ML classification algorithms has proven to achieve greater precise performance compared to utilizing a solitary technique (Rajab *et al*., 2024). The study employed bagging, stacking, soft voting and adaboost.

Bagging works by reducing variance and preventing overfitting by training multiple models on different subsets of the training data and averaging their predictions (Rajab *et al*., 2024). RF was preferred for the bagging method as it uses multiple decision trees during training (Rajab *et al*., 2024). Training of the bagging model was done using the randomForest function in R.

Stacking uses basic-level meta classifiers and amalgamates them with meta-learner classifiers (Rajab *et al*., 2024). The base learners used for stacking are RF, DT, KNN, GB, NB and XGBoost and LR as the base classifier. Prediction models were first obtained from each of the base models. A new dataset (predictions_stacked) was created and it contained all base learners and the actual target variable (Diagnosis).

Soft Voting used LR, RF, DT, KNN, GB, NB and XGBoost as base models. These base models were averaged and the predicted probabilities were combined. AdaBoost used the ada package in R, with 100 iteration per iter. AdaBoost adjusts the weights of misclassified instances to improve the performance of weak learners.

### Individual and ensemble model performance evaluation

The individual and ensemble models were evaluated on their performance in malaria prediction using metrics such as accuracy, recall, precision, confusion matrix, F1 score and AUC-ROC. Predictions and predicted probabilities were generated for each model and the performance metrics were calculated and compared. For ensemble models, the same metrics were used to assess the effectiveness. Model performance was also compared between individual and ensemble models.

### Ethical review

Ethical approval was obtained from Gweru Provincial Hospital’s Research Ethics Committee and Gutu Rural District Hospital’s Research Ethics Committee. Measures were implemented to ensure any ethical issues relating to the study were addressed. All patient identifiers such as names, national identification number and contact details were removed during data collection. For issues to do with data security, the dataset was kept secure and accessible to authorized people only.

## Results

### Analysis of demographic and clinical variables associated with malaria diagnosis

A total of 637 participants with an age range of 1 to 90 years were included in this study. The mean age was 30.7 ± 21.4 years, indicating a wide variation in participant ages. Out of the 637 participants 49.9% were from Gweru Provincial Hospital while 50.1% were from Gutu Mission Hospital. The distribution of gender was similar between groups, with no significant difference observed in their diagnosis results [p = 0.483, Table 2].

**Table 2:** Baseline characteristics for all the participants in this study. *Percentages shown in the table are column percentages (within Diagnosis groups). Significant p-values indicated by * (<0.05), ** (<0.01)*.

| Variable | Category | Negative<br>(n=562) | Positive<br>(n=75) | Total<br>(n=637) | p-value |
| --- | --- | --- | --- | --- | --- |
| **Gender** | Male | 283 (50.4%) | 41 (54.7%) | 324 (50.9%) | 0.483 |
|  | Female | 279 (49.6%) | 34 (45.3%) | 313 (49.1%) |  |
| **Fever** | Yes | 371 (66.0%) | 63 (84.0%) | 434 (68.1%) | 0.002 ** |
|  | No | 191 (34.0%) | 12 (16.0%) | 203 (31.9%) |  |
| **Chills** | Yes | 350 (62.3%) | 61 (81.3%) | 411 (64.5%) | 0.001 ** |
|  | No | 212 (37.7%) | 14 (18.7%) | 226 (35.5%) |  |
| **Headache** | Yes | 521 (92.7%) | 69 (92.0%) | 590 (92.6%) | 0.826 |
|  | No | 41 (7.3%) | 6 (8.0%) | 47 (7.4%) |  |
| **Diarrhea** | Yes | 149 (26.5%) | 31 (41.3%) | 180 (28.3%) | 0.007 ** |
|  | No | 413 (73.5%) | 44 (58.7%) | 457 (71.7%) |  |
| **Abdominal Pain** | Yes | 166 (29.5%) | 38 (50.7%) | 204 (32.0%) | <0.001 ** |
|  | No | 396 (70.5%) | 37 (49.3%) | 433 (68.0%) |  |
| **Travel History** | Yes | 198 (35.2%) | 37 (49.3%) | 235 (36.9%) | 0.017 * |
|  | No | 364 (64.8%) | 38 (50.7%) | 402 (63.1%) |  |
| **Location** | Rural | 287 (51.1%) | 49 (65.3%) | 336 (52.7%) | 0.020 * |
|  | Urban | 275 (48.9%) | 26 (34.7%) | 301 (47.3%) |  |
| **Age Group** | 0-5 | 92 (16.4%) | 8 (10.7%) | 100 (15.7%) | 0.298 |
|  | 6-15 | 62 (11.0%) | 6 (8.0%) | 68 (10.7%) |  |
|  | 16-30 | 160 (28.5%) | 19 (25.3%) | 179 (28.1%) |  |
|  | 31-45 | 104 (18.5%) | 21 (28.0%) | 125 (19.6%) |  |
|  | 46-60 | 89 (15.8%) | 11 (14.7%) | 100 (15.7%) |  |
|  | >60 | 55 (9.8%) | 10 (13.3%) | 65 (10.2%) |  |

However, participants who tested positive were significantly more likely to report fever (84.0% vs. 66.0%, p = 0.002), chills (81.3% vs. 62.3%, p = 0.001), diarrhea (41.3% vs. 26.5%, p = 0.007), and abdominal pain (50.7% vs. 29.5%, p < 0.001) compared to those who tested negative [Table 2]. A higher proportion of positive cases had a recent travel history (49.3% vs. 35.2%, p = 0.017) and resided in rural areas (65.3% vs. 51.1%, p = 0.020). No significant differences were observed in the distribution of headache or age groups between the two diagnosis categories [Table 2].

### Feature selection

A correlation analysis of selected numeric and binary-encoded variables (age, gender, headache, fever, abdominal pain, diarrhea, chills and travel history) confirmed the predictors are relatively independent of each other. The pairwise Pearson correlation coefficients [Figure 2] showed most of the correlations are close to zero, an indication of weak linear relationships between variables. The highest observed correlation was between chills and fever, with a coefficient of -0.40. Travel history showed weak to negligible correlations with all other features, except a moderate negative correlation with age ( r=−0.08).

**Figure 2:**
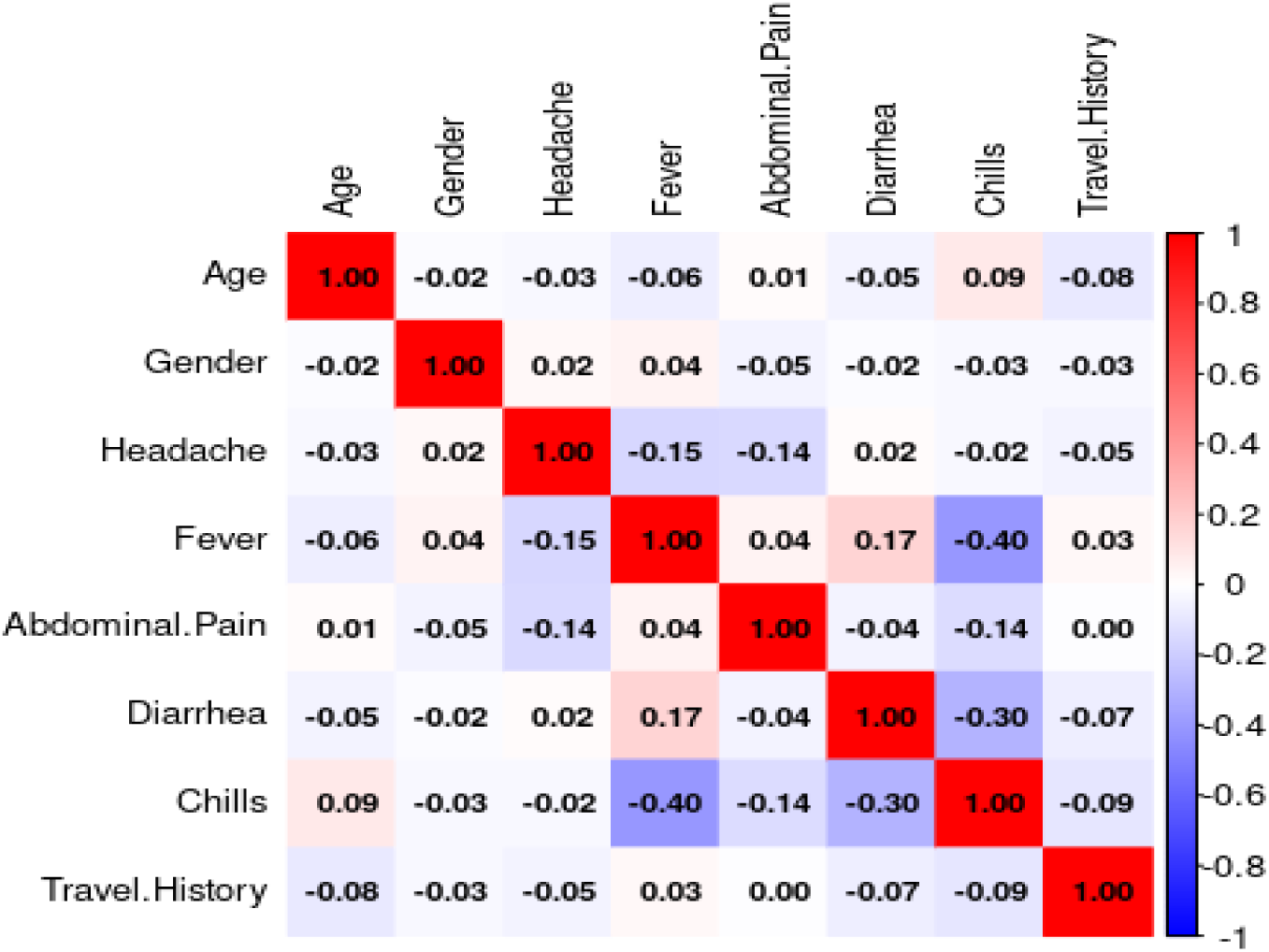
Feature Correlation Matrix for Malaria Predictors

VIF values for the predictors were all below 5, indicating no significant collinearity among predictors [Table 3]. The highest VIF was 1.35 for chills and the lowest VIF was 1.01 for gender. Findings align with the weak correlations observed in the correlation matrix [Figure 2].

**Table 3:** VIF analysis output for correlation analysis

| Variable | VIF Value |
| --- | --- |
| Chills | 1.35 |
| Fever | 1.24 |
| Diarrhoea | 1.12 |
| Headache | 1.06 |
| Abdominal Pain | 1.06 |
| Residence | 1.04 |
| Travel History | 1.04 |
| Age | 1.03 |
| Gender | 1.01 |

The top five identified variables [Figure 3] using RFE were chills, fever, travel history and abdominal pain, all of which showed statistical significance with chi square values [Table 2], chills: p=0.0019), fever: p=0.003, diarrhea: p=0.01, travel history: p=0.02 and abdominal pain: p=0.0004.

**Figure 3:**
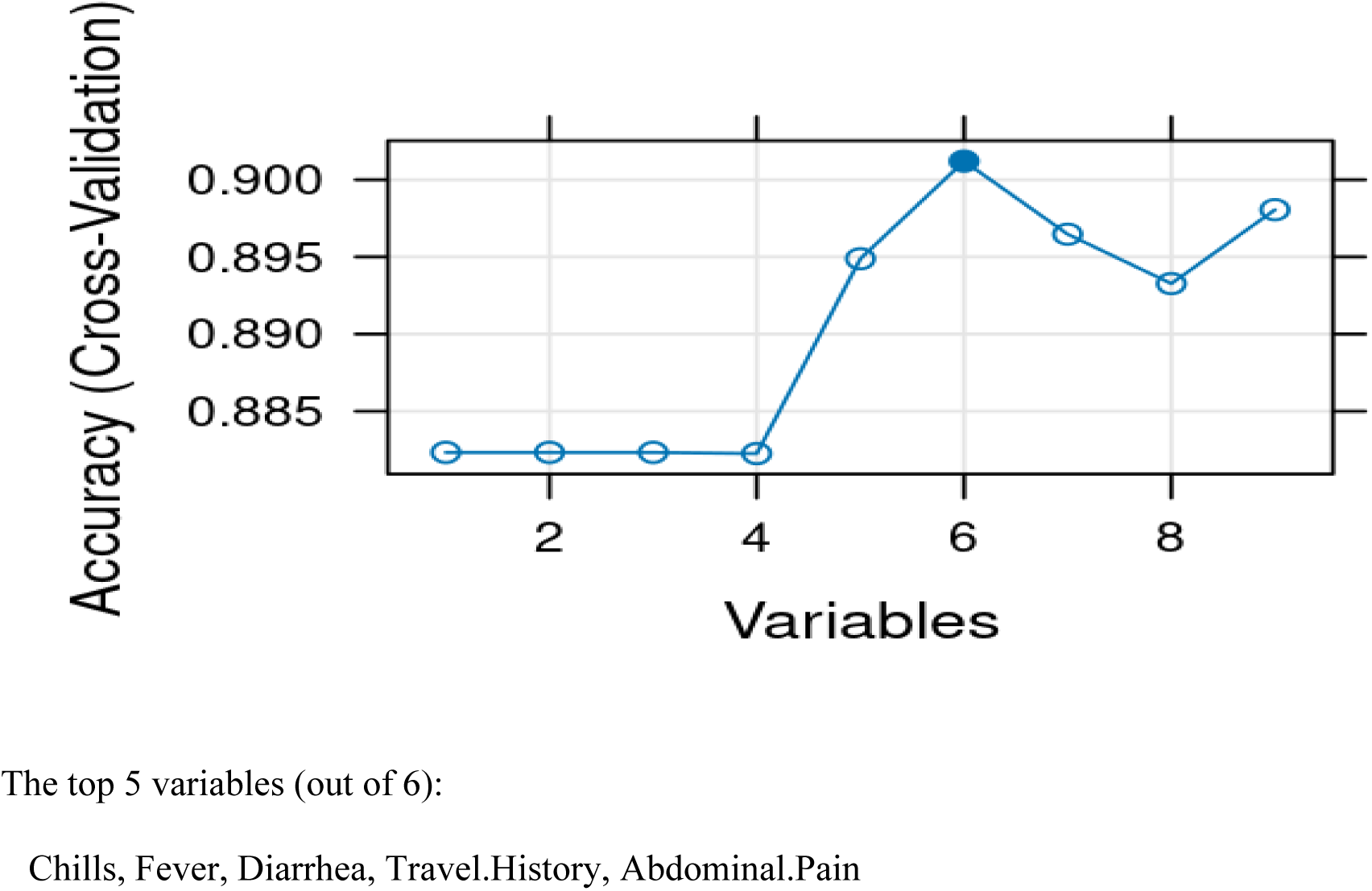
Feature Importance Analysis with Recursive Feature Elimination

SMOTE successfully addressed the data imbalance issue to improve model performance [Figure 4].

**Figure 4:**
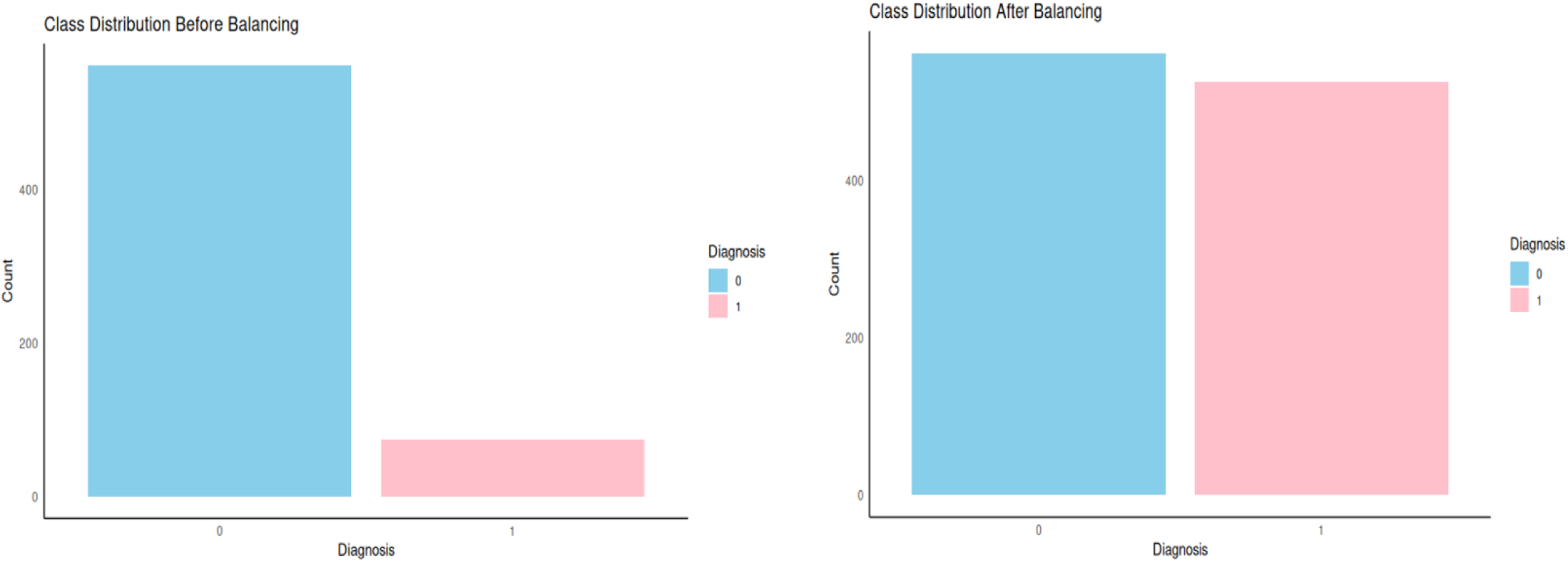
Class distribution before and after balancing with SMOTE

### Machine learning

The performance of the seven ML individual classifiers (LR, RF, DT, GB, KNN, NB and XGBoost) were evaluated using six metrics. XGBoost got the highest accuracy (0.95). GB and RF both got an accuracy of 0.94. DT achieved an accuracy of 0.89, while LR had 0.83 and NB had 0.82, showing lower but comparable performance. KNN had the lowest accuracy of 0.69. XGBoost achieved the highest precision (0.93). RF and GB had precision values of 0.91 and 0.92 respectively. DT (0.87) and LR (0.85) achieved moderate precision values. KNN had the lowest precision of 0.68. Recall was strongest for RF at 0.99. XGBoost and GB both got a recall 0.98. DT obtained 0.93 showing a good recall performance. LR (0.82) and NB (0.77) demonstrated more limited sensitivity. KNN achieved 0.79 recall, performing better in this metric than in others.[Figure 5].

**Figure 5:**
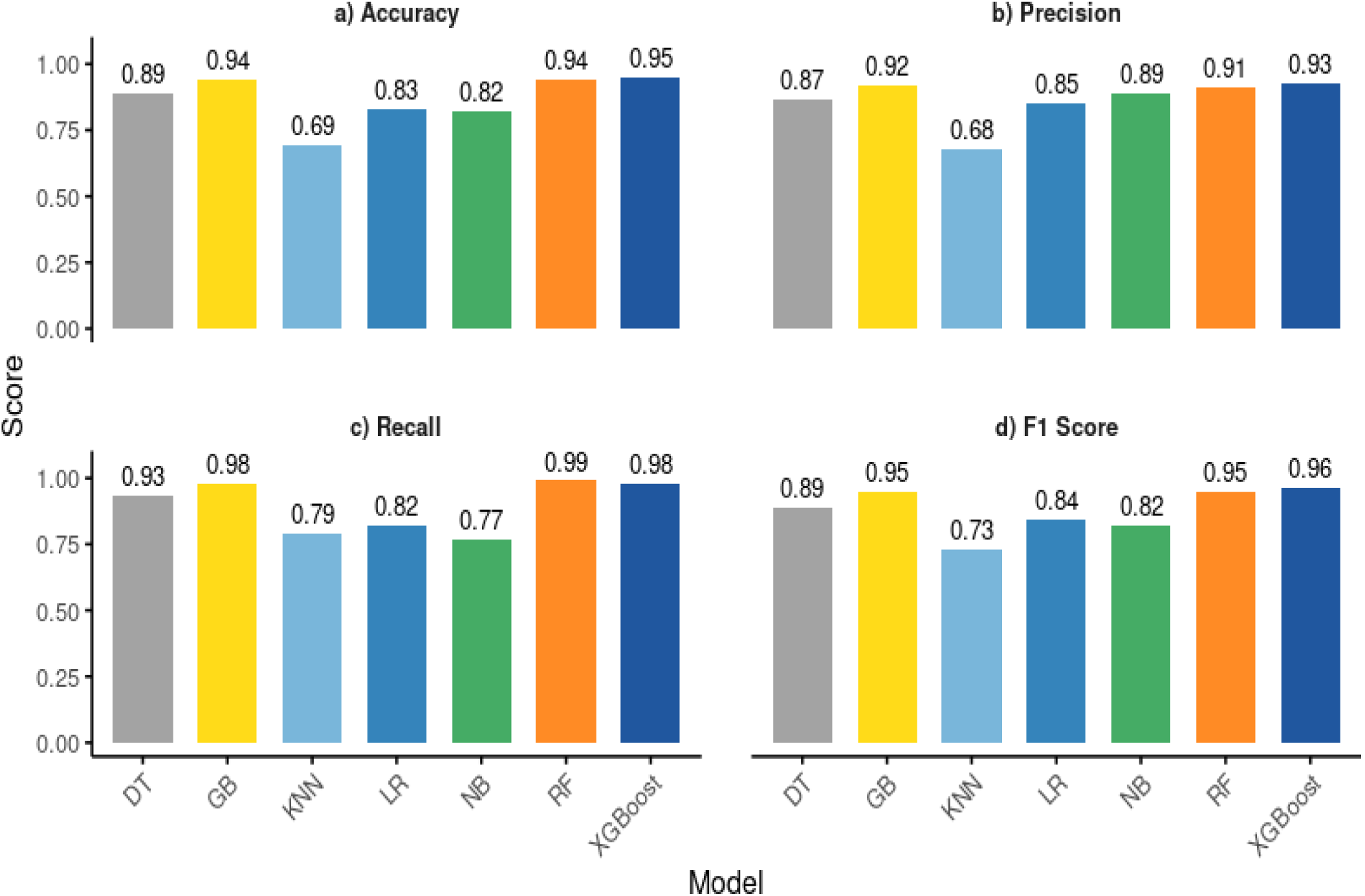
Model Performance Evaluation on Test set

The AUC-ROC values, indicating overall classification ability across all thresholds, were strongest for XGBoost and GB (both 0.99). RF achieved 0.98, DT (0.95) and NB (0.90), while LR achieved 0.89. KNN (0.75) had the lowest AUC-ROC performance [Figure 6].

**Figure 6:**
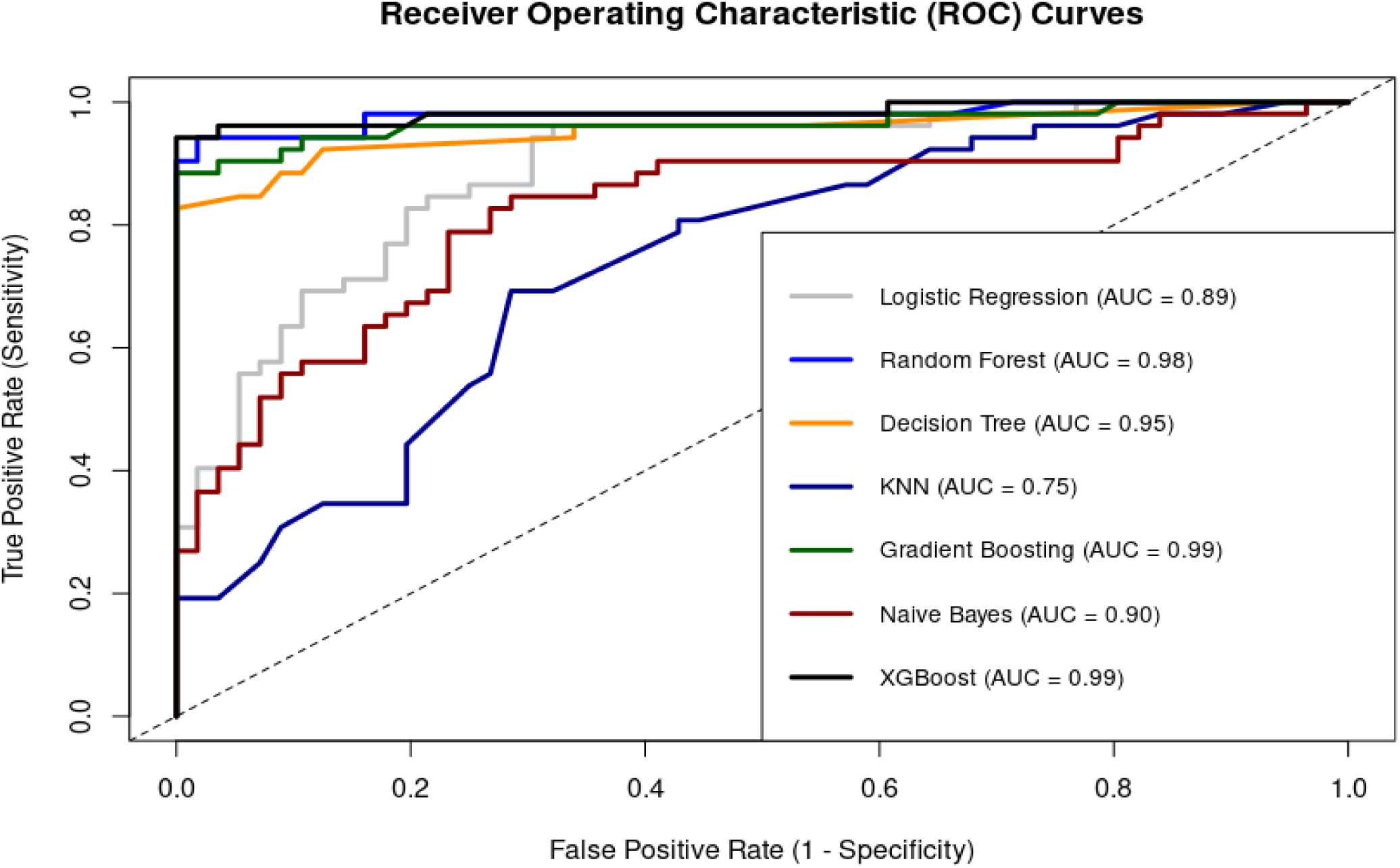
Receiver Operating Characteristic Curve Analysis on test set Performance

Confusion matrices were used to evaluate the models, where each matrix shows the number of actual malaria cases (positives) and non-cases (negatives) correctly or incorrectly predicted by the model [Figure 7]. LR correctly classified 40.7% true positives (TP) and 42.6% true negatives (TN), then misclassified 7.4% of actual positives as false negatives (FN) and 9.3% of actual negatives as false positives (FP). XGBoost achieved strong performance, with 44.4% TP and 51.9% TN. Impressively, it recorded 3.7% FN and 0% FP, making it the model with the lowest error. GB correctly identified 43.5% of TP and 50.9% of TN. It had minimal misclassifications of 4.6% FN and only 0.9% FP.

**Figure 7:**
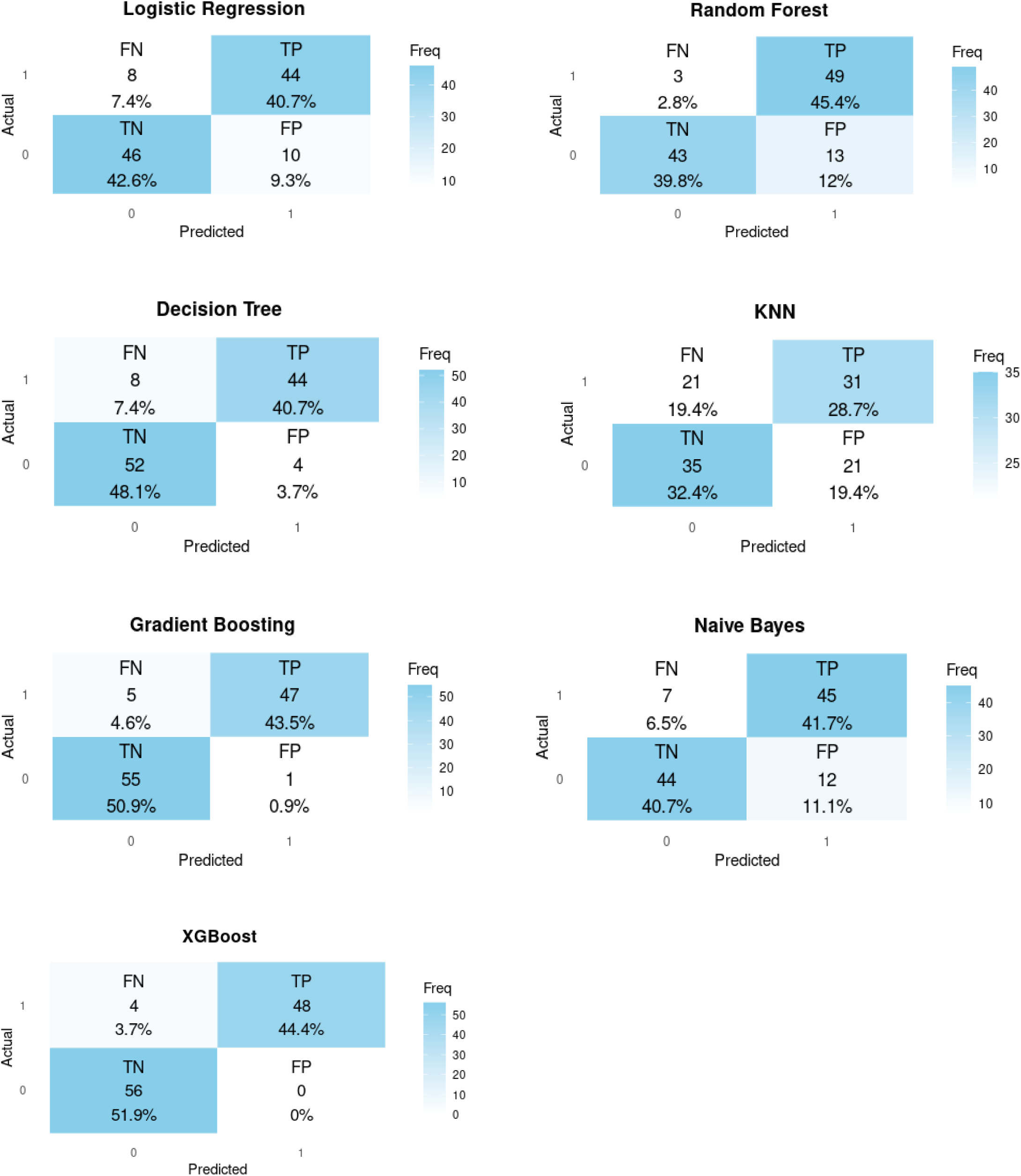
Confusion Matrix Performance for Models on Test set (1= Positive, 0= negative)

Comparative performance analysis of models between the test and evaluation sets showed consistent patterns in predictive accuracy and reliability. XGBoost showed superior performance across all metrics, achieving the highest accuracy (0.95 test, 0.93 eval), F1-score (0.96 test, 0.93 eval), AUC-ROC (0.99), precision (0.93) and recall (0.98). RF and GB models had a nearly similar performance to XGBoost. DT showed moderate performance with accuracy and F1-scores around 0.89 [Figure 8].

**Figure 8:**
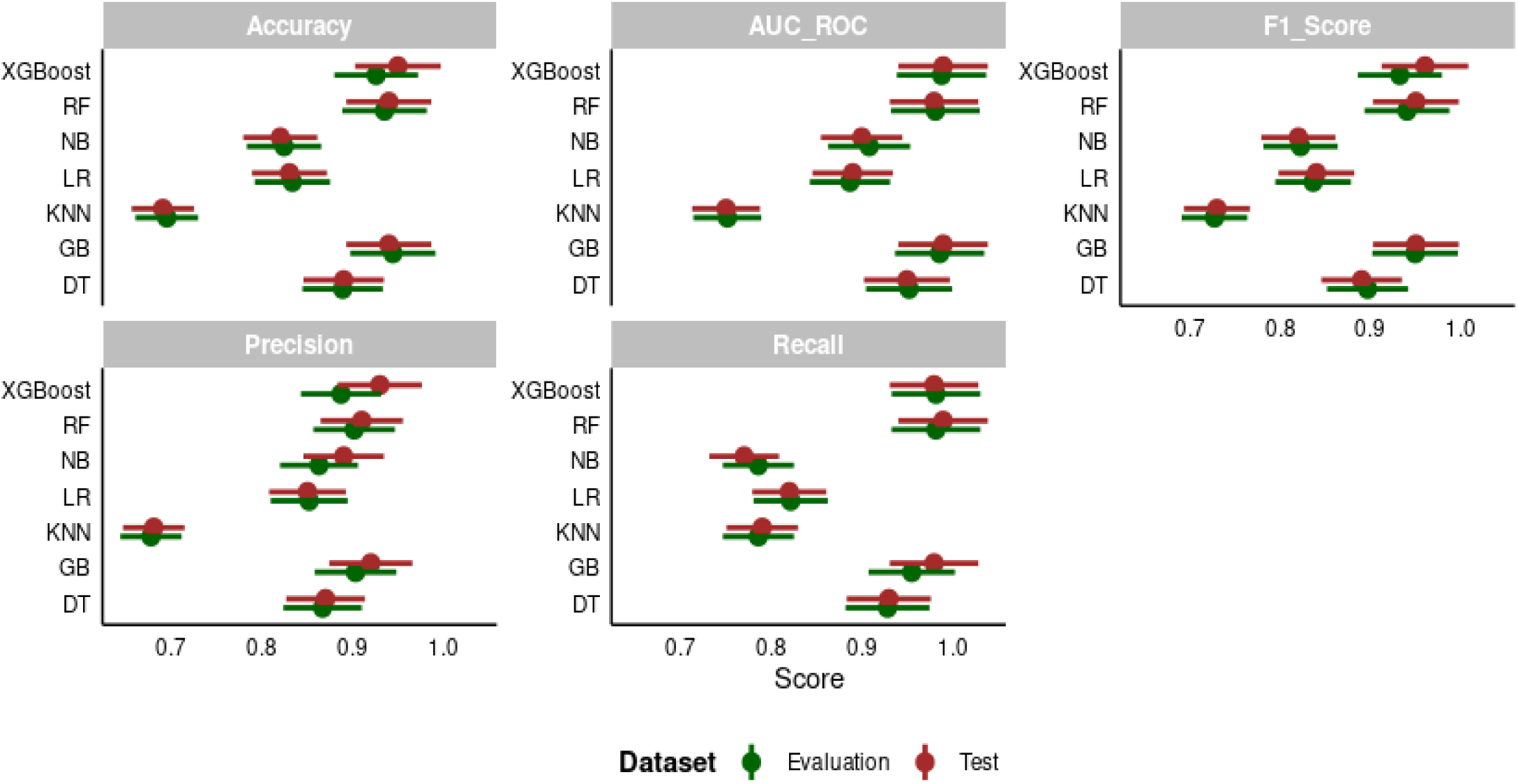
Comparison of Model Performance on evaluation and Test Sets.

The Stacking model demonstrated the highest overall performance across all evaluation metrics. Stacking has an accuracy of 96% while Soft Voting, Bagging and AdaBoost classifiers achieved accuracy score of 94%. Soft Voting had a slightly higher precision (93%). Stacking had the highest precision of 95% [Figure 9].

**Figure 9:**
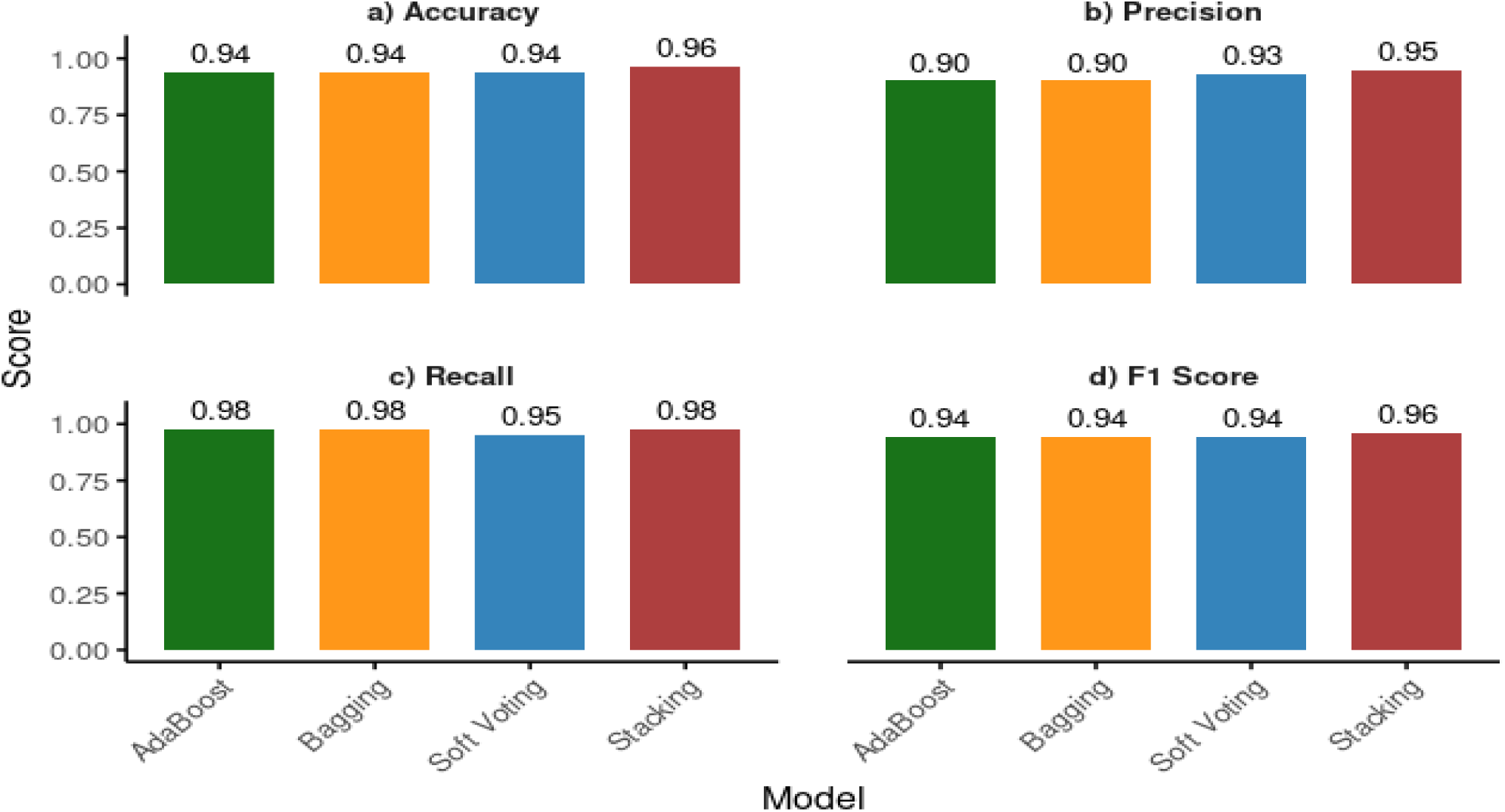
Ensemble Model Performance on test set

The ROC curves showed exceptional discriminative performance across all ensemble models. Soft Voting and Bagging achieved a high classification capability (AUC = 0.99). AdaBoost and Stacking had AUC values of 0.98. All ensemble models maintained high sensitivity with their curves hugging the top-left corner of the plot, a nature only high-quality classifiers take. The clustering of AUC scores between 0.98 and 0.99 confirms that all ensemble methods provide clinically reliable diagnostic performance [Figure 10].

**Figure 10:**
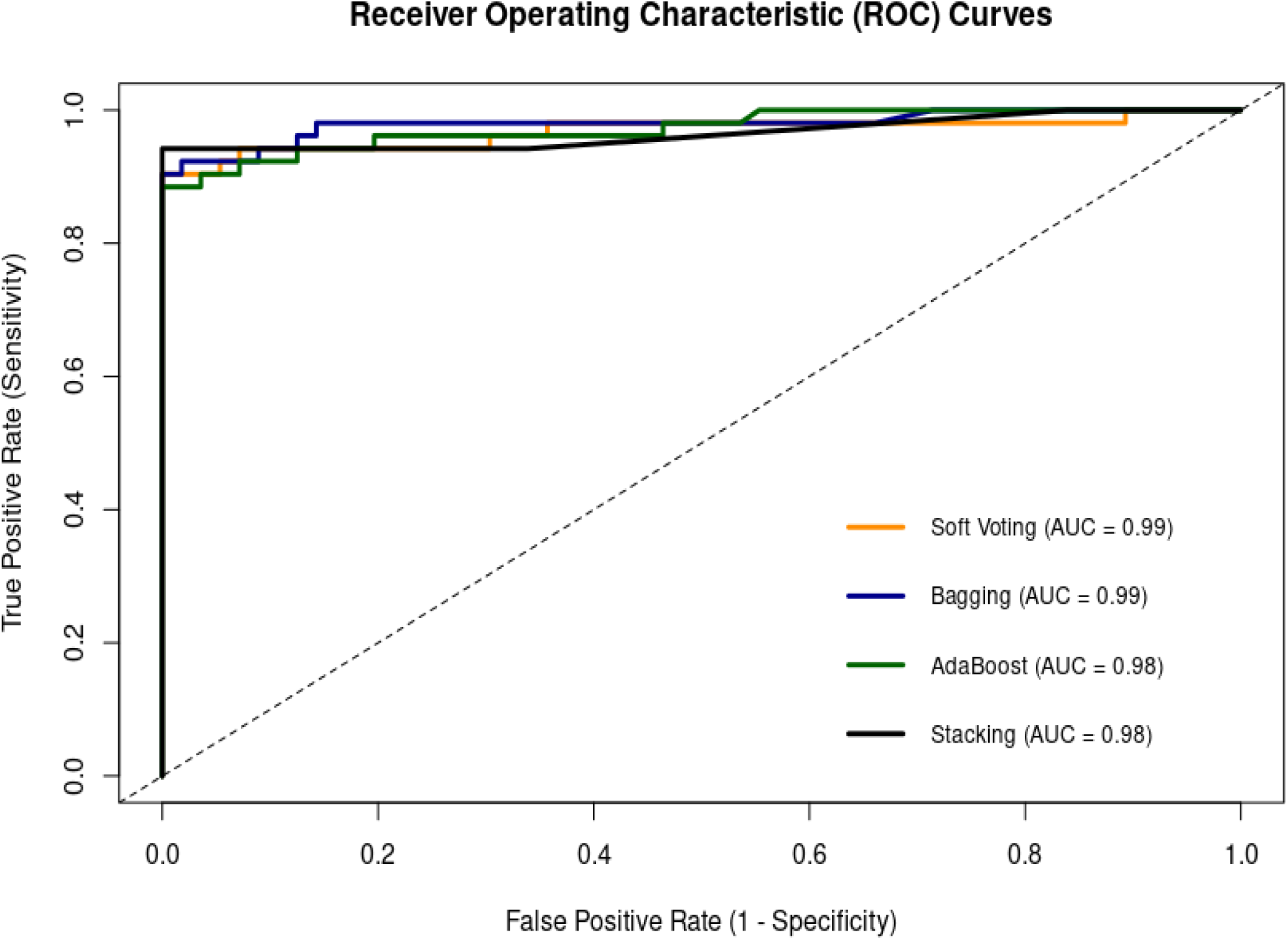
ROC Curve Analysis on test set Performance for Ensemble Models

Confusion matrices for the four ensemble models show distinct performance quality in malaria classification [Figure 11]. Soft Voting correctly identified 48 TP cases (44.9%) and 53 TN (49.1%), with error rates 4 FN (3.7%) and 3 FP (2.8%). Bagging had stronger specificity, with only 1 FP and 55 TN, with 6 FN at 5.6%. AdaBoost demonstrated the best sensitivity among all ensemble models, with 3 FN (2.8%) and 49 TP (45.4%) and only 1 FP. Stacking matched AdaBoost’s sensitivity with 3 FN at 2.8%, 49 TP at 45.4%, but with more FP. All models maintained FP rates below 3%.

**Figure 11:**
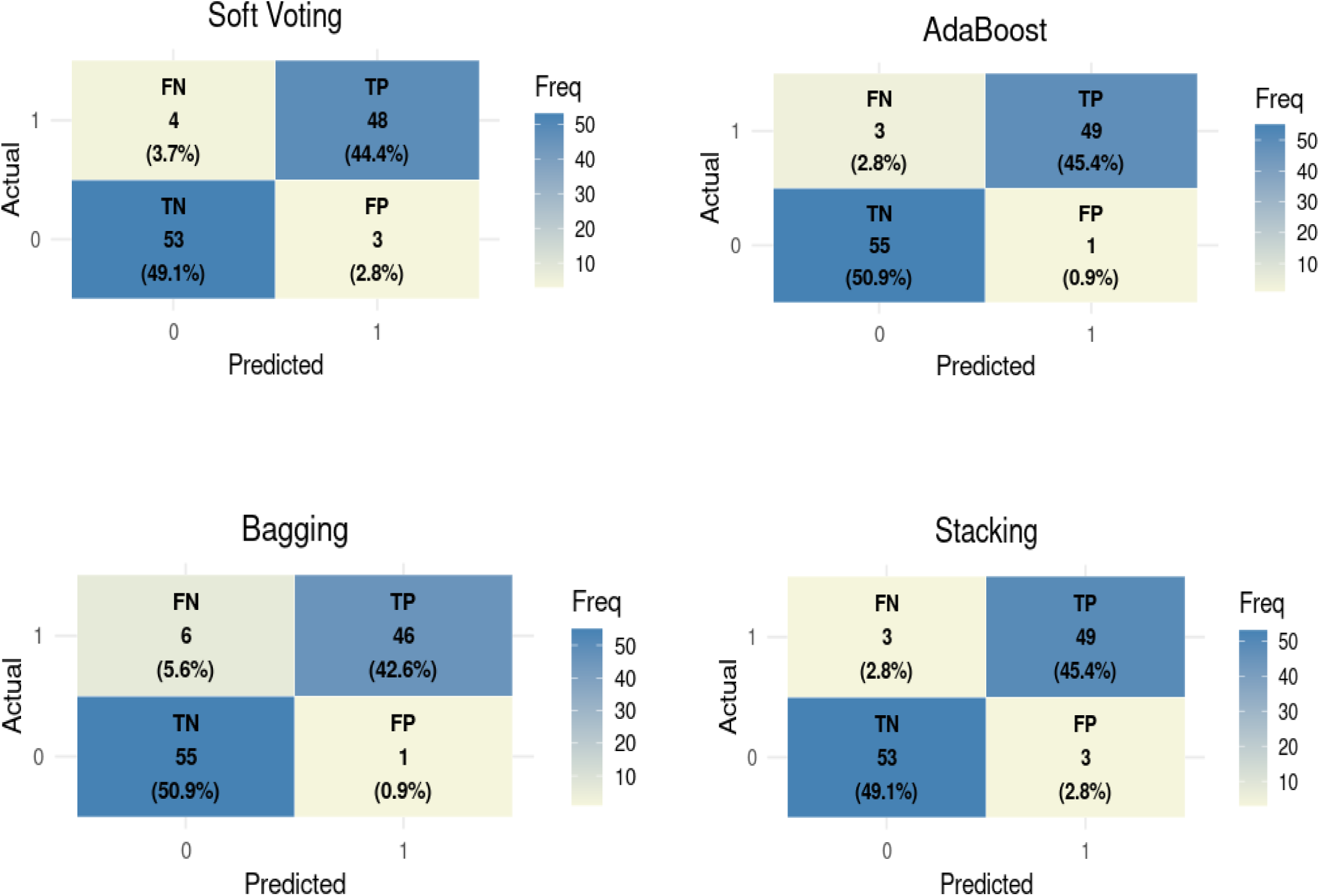
Confusion Matrix Performance for Ensemble Models on Test set (1= Positive, 0= negative)

## Discussion

The findings of this research demonstrates the potential impact of ML ensemble models in enhancing malaria diagnosis particularly in developing countries where there is poor accessibility and insufficient diagnostic tools for malaria. Our results show that the Stacking ensemble model outperformed all individual models, obtaining accuracy = 0.96, precision = 0.95, recall = 0.98, F1 score = 0.96 and AUC-ROC = 0.98.

Of the 637 participants from Gweru Provincial Hospital (49.9%) and Gutu Mission Hospital (50.1%), a proportion of 50.9 were males and 49.1% were females. The age of the participants had a mean of 31 ± 21 years. A total of 75 cases were positive, with 54.7% of the 75 cases being males and 45.3% females showing a slightly higher prevalence in males, although the difference did not attribute any statistical significance (p= 0.56). Clinical symptoms such as chills, fever, abdominal pain and diarrhea were significant predictors of malaria (p < 0.05) highlighting the dependence of clinical symptoms as diagnostic markers for malaria. While most demographic features were not statistically significant predictors of malaria, travel history proved to have a significant effect towards malaria incidence as individuals who travel may have different risk exposure and prevention behaviours. These results align with existing literature which emphasize how physical clinical symptoms of malaria can be relied on for diagnosis, especially in malaria endemic areas (Atukunda *et al*., 2021). Some studies argue that demographic factors are as important as clinical symptoms, reporting gender and age as having a huge impact in malaria incidence (Doreswamy and Al-Sudani, 2022; Lynch *et al*., 2015). Other literature report that while demographics are found to be related to malaria incidence, different geographical and socio-economic conditions impact their significance to malaria epidemiology (Ekusai-Sebatta *et al*., 2021; Brungard *et al*., 2015). Early diagnosis of malaria through timely identification of clinical symptoms can potentially improve malaria management and control in endemic localities (Arinaitwe *et al*., 2018; Doreswamy and Al-Sudani, 2022).

The performance evaluation of the ML models measured by considering all the metrics showed that ensemble models outperform individual ML models. XGBoost achieved an accuracy of 0.95 while RF and GB got 0.94 indicating strong generalizability. A comparison on the individual models’ performance on the evaluation and test datasets [Figure 8], showed consistent performance proving that there was minimal overfitting due to effective hyperparameter tuning. Models such as KNN and NB which demonstrated moderate model performance could suggest their applicability for specific scenarios with fewer complexities.

While all the ML ensemble models achieved high diagnostic performance [Figure 9], the Stacking ensemble model outperformed the other ensemble models obtaining a high malaria diagnostic performance across all metrics. Bagging, Stacking and AdaBoost achieved 0.98 recall, and Soft Voting got 0.95. High recall in malaria diagnosis means reliable identification of positive cases reducing risk of severe complications. Mahajan *et al*. (2024) and Solimo *et al*. (2024) obtained very high precision scores for Bagging, Stacking and AdaBoost, emphasizing that ensemble models are reliable in boosting sensitivity in malaria diagnosis. F1 scores for the ensemble models were high, with Stacking and AdaBoot at 0.96. Soft Voting and Bagging got 0.94 indicating a strong overall performance. The study’s results for the F1 score of the ensemble model performance highlight the potential of the ensemble models at balancing precision and recall. Ensemble models demonstrated exceptional reliability in making predictions with minimal errors [Figure 11]. The top performing individual models in our study were XGBoost, GB and RF while the top performing ensemble model was stacking.

## Conclusion

Our investigation demonstrated that using clinical and demographic data for malaria diagnosis with ensemble ML models is viable. These ensemble models offer a scalable and cost effective alternative or supporting tool to traditional diagnostic methods of malaria in resource limited settings. Clinical symptoms such as chills, fever and abdominal pain demonstrated strong predictive power, highlighting their importance and usability in symptom-based screening tools. These findings can assist in malaria control programs in high burden areas.

Notably, limitations such as sample size, data diversity, model complexity and possible deployment issues were are acknowledged. Integrating these stacking ensemble models into a user-friendly digital platform can help health workers gain access to reliable diagnostic support reducing burden on health systems. Validating the proposed models in real time clinical workflows should be adopted to assess their clinical applicability. Model integration should be piloted into hospital systems or mobile health platforms to assess clinical acceptance and usability.

This study contributes to the growing evidence of supporting the use of AI in global health. By demonstrating the feasibility of ML driven malaria prediction, this study lays a foundation for scalable and tailored data informed diagnostic tools that can be used in supporting infectious disease management in resource limited settings. It also adds to the countless efforts of developing nations towards the achievement of sustainable development in the healthcare sector under the SDG-3 pillar of the United Nations.

## Future Works

Future research includes focusing on large scale validation and real world implementation to ensure model’s effectiveness in clinical settings. To further advance the performance of ensemble models focusing more on investigating the impact of more than base models within ensemble models and combining ensemble models to explore their impact.

## Data Availability

Yes, the data is available.

## List of Abbreviations

ML: Machine Learning
LR: Logistic Regression
RF: Random Forest
DT: Decision Trees
GB: Gradient Boosting
KNN: K-Nearest Neighbors
NB: Naive Bayes
AUC-ROC: Area Under the Receiver Operating Characteristic Curve
SMOTE: Synthetic Minority Oversampling Technique

## Declarations

### Ethics Approval and Consent to Participate

The study was approved by the Gweru Provincial Hospital’s Research Ethics Committee and Gutu Rural Research Ethics Committee. Patient records were anonymized, and informed consent was obtained from all participants.

### Consent for Publication

Not applicable.

### Availability of Data and Materials

The datasets used and/or analyzed during the current study are available from the corresponding author on reasonable request.

## Authors’ contributions

### Acknowledgements

We acknowledge Gweru Provincial Hospital and Gutu Mission Hospital for approving and allowing us to carry out this study using the patient data collected from their medical records.

